# Efficacy and Safety of Ayurveda Intervention (AYUSH 64) as add-on therapy for patients with COVID-19 infections – An open labelled, Parallel Group, Randomized controlled clinical trial

**DOI:** 10.1101/2021.08.10.21261836

**Authors:** Pankaj bhardwaj, Pawan Kumar Godatwar, Jaykaran Charan, Sanjeev Sharma, Shazia Shafi, Nishant Chauhan, Pratibha Vyas, Naveen Dutt, Naresh Midha, Ramniwas Jalandra, Meenakshi Sharma, Vijaya Lakshmi Nag, Suman Sharma, Sarvesh Kumar Singh, Praveen Sharma, Sanjeev Misra

## Abstract

COVID-19 pandemic impacted human health and the global economy. There is a huge uncertainty about the management of this disease, many drugs including some older drugs are being tested for efficacy and safety including the medicines from the complementary and alternative system. The Central Council for Research in Ayurvedic Sciences, India’s apex body for Ayurvedic research and development under the Ministry of AYUSH, has developed a poly-herbal drug called AYUSH 64 for covid 19 which is having role in the COVID-19. This study was designed with the aim of assessing the efficacy and safety of AYUSH 64 in mild covid-19 patients as add on therapy with standard treatment.

It was an open labelled, comparative, parallel group, Randomized controlled clinical trial. Total 60 stage I (mild) COVID 19 positive subjects were recruited, 30 were assigned to AYUSH 64 as an add on therapy along with the standard treatment and 30 were assigned to standard treatment as per the protocols. RT-PCR test was done as per government guidelines and protocol. Along with the RT-PCR clinical laboratory tests were also performed at screening as well as on the discharge as per the study schedule.

Absolute events of negative RT-PCR at day 5 were more in the AYUSH 64 group as compared to control group though not statistically significant (70% Vs 54%, p=0.28) is a clinically important finding. There was an equivalence between AYUSH 64 and control group for fever and respiratory symptoms or important lab parameters. No serious adverse event was reported from any group.

AYUSH 64 is safe and has a beneficial effect though not statistically significant as compared to control group, this may be because of the less sample size which need to be confirmed by studies with large sample size.

**Trial Registration:** Clinical Trial Registry of India - CTRI/2020/06/026002

## Introduction

COVID-19 has emerged as the latest pandemic affecting millions of the people worldwide. Coronavirus disease (COVID-19) is an infectious disease caused by the SARS-CoV-2 virus. [1] World Health Organization (WHO) labelled it a Public Health Emergency of International Concern (PHEIC) on January 30, 2020, and a global pandemic on March 11, 2020. [2] Coronavirus disease is transmitted by droplets through inhalation or physical contact with the affected patient. SARS-CoV-2 is a zoonotic virus in the Coronaviridae family that can infect humans and a variety of animals. SARS-CoV-2 is believed to have been transmitted to humans by an unidentified intermediary species and then spread from human to human. [2] Looking at the morbidity and mortality associated with this disease, there is a urgent need of effective therapy for the treatment of this disease but till now no effective therapy is available.

The lack of approved effective drug therapeutic protocols for COVID-19 will make treating newly emerged COVID-19 infections around the world difficult. Not only the drug repurposing but alternative medicine system should be explored to find cure for this disease which is associated with such a high number of deaths considering overall exposed population. [3,4]

The central Council for Research in Ayurvedic sciences has developed a poly-herbal drug “Ayush - 64” whose constituents is found to be effective in various diseases having viral and parasitic origin. [5-19] The composition of AYUSH 64 includes aqueous extract of Saptaparna 100 mg (Alstonia scholaris), Katuki 100 mg (Picrorhiza kurroa), Kiratatikta 100 mg (Swertia chirata) and powder of Kuberaksha 200 mg (Caesalpinia crista) in the ratio of 1:1:1:2. As per the in-silico study based on docking, it was found that 36 chemical compounds of the AYUSH 64 have good potential for anti SARS COV-2 activity. [20] It was also found to be effective in Influenza like Illness (ILI) in a clinical study. [21] It was worthwhile to explore the efficacy and safety of this formulation for COVID-19, considering its good efficacy in influenza like disease. Hence this study was designed with the aim of assessing the AYUSH 64 in addition to the standard treatment in mild COVID-19 patients in addition to the standard treatment for the efficacy and safety.

## Material and methods

### Study design

It was an open labelled, parallel group, prospective, Randomized clinical trial.

### Study objectives

The primary objective of this study was to assess the efficacy of AYUSH - 64 in the mild COVID-19 patients based on negative RT-PCR on 5^th^ day of treatment. Secondary objectives were to assess the efficacy of the AYUSH 64 based on clinical and laboratory parameters and to evaluate the safety of AYUSH 64 in these patients based on hematological parameters and adverse events.

### Subject selection

The study was conducted in AIIMS Jodhpur on stage I (mild) COVID 19 positive patients. A total of 60 subjects were recruited. Based on the computer based randomized sequence 30 subjects were enrolled in group 1 (intervention arm) and 30 in group 2 (control arm). Subjects aged 18 to 60 years, who tested RT-PCR positive for COVID-19 and were categorized under stage I- mild (early infection), willing to take medicines orally and to provide signed informed consent were included in the study. Pregnant, Lactating women, patients with CKD (chronic kidney disease), and those not willing to participate were excluded from the study. Subjects with severe COVID-19 or acute respiratory distress syndrome or those hospitalized, on invasive mechanical ventilation or extracorporeal membrane oxygenation, with alanine transaminase (ALT) or aspartate transaminase (AST) more than two times the upper limit of normal, were excluded.

### Therapeutic intervention

Group 1 (Intervention Group): The intervention arm was given AYUSH 64, a polyherbal drug developed by Central Council of Research in Ayurveda Science (manufactured by Unijiles life sciences LTD., Nagpur, India). It was given in the dose of 2 tablets (500 mg each) thrice daily i.e. 3 gm /day orally after food along-with water for 07 days as an add-on treatment to standard care.

Group 2 (Control Group): The control group was given standard treatment as per the guideline of the Ministry of Health and Family Welfare, Government of India.

### Study procedure

This study was started after registering trial in Clinical Trial Registry of India (CTRI) (CTRI/2020/06/026002) and getting permission from the Institutional Ethics Committee (AIIMS/IEC/2020-21/3036). The subjects were fully informed about the aims, procedures, discomforts and expected benefits of the trial by the principal investigator. COVID-19 patient diagnosed by RT-PCR who came for the treatment at the AIIMS Jodhpur were considered for inclusion. All eligible patients were randomized into the intervention and control group by computer generated randomization sequence. Allocation concealment was done centrally through telephone. Group 1 were give Ayush 64 as add on therapy along with the standard treatment for Covid patients as per the guidelines, and Group 2 were given local standard care of treatment as per the protocols. These patients were followed up for RT-PCR testing and blood parameters. RT-PCR was done on the 5th day. Repeat RT-PCR was done as per the standard protocol if needed.

Laboratory tests were performed at screening as well as on the discharge as per the study schedule which include CBC, LFT, RFT, RBS, Hs-CRP, LDH, S. Ferritin, D-dimer and IL-6. Clinical parameters including symptoms and signs were assessed daily as per the standard of care. During the treatment each subject were having access to the investigators which included AYUSH practitioner also.

### Statistical analysis

Descriptive statistics was reported in the form of frequency, percentages, mean and standard deviation. Difference in frequency of negative RT-PCR on 5th day was analyzed by Fisher’s exact test and blood parameters were compared by unpaired t test. Statistics for Windows, version 23 (SPSS Inc., Chicago, IL, USA) was used for the analysis.

## Results

All 60 subjects (30 in the intervention arm and 30 in the control arm) completed the follow up of five days. **Table 1** depicts the sociodemographic profile of the subjects enrolled in this study. There was no significant difference in any sociodemographic parameters between both groups.

**Table 1:** sociodemographic profile of the subjects.

|  | Group 1<br>(Ayush- 64)<br>(n = 30, n %) | Group 2 (Control)<br>(n = 30, n %) |
| --- | --- | --- |
| **Gender** |  |  |
| Female | 1 (3.3) | 1(3.3) |
| Male | 29 (96.6 ) | 29 (96.6 ) |
| **Marital status** |  |  |
| Married | 22 (73.33) | 23 (76.67) |
| Unmarried | 8 (26.67) | 7 (23.33) |
| **Educational status** |  |  |
| Illiterate | 5 (16.67) | 2 (6.66) |
| Upto 10<sup>th</sup> | 9 (30) | 8 (26.67) |
| Upto 12<sup>th</sup> | 2 (6.67) | 7 (23.33) |
| Graduate | 12 (40) | 10 (33.33) |
| Post graduate | 2 (6.67) | 3 (10) |
| **Occupation** |  |  |
| Desk work | 3 (10) | 4 (13.33) |
| Field work with physical labour | 19 (63.33) | 14 (46.67) |
| Field work | 4 (13.33) | 4 (13.33) |
| House Wife | 1 (3.33) | 0 (0) |
| Student | 2 (6.67) | 3 (10) |
| Others | 1 (3.33) | 5 (16.67) |
| **Socio-economic status** |  |  |
| Above Poverty line | 25 (83.33) | 28 (16.66) |
| Below Poverty line | 5 (16.66)) | 2 (6.06) |
| **Habitat** |  |  |
| Urban | 26 (86.66) | 30 (100) |
| Semi-urban | 3 (10) | 0 (0) |
| Rural | 1 (3.33) | 0 (0) |

On comparing both the groups for frequency of RT-PCR negative subjects on day 5^th^, it was found that 21 (70%) subjects from the AYUSH 64 group and 16 (54%) subjects from control group were RT-PCR negative on 5^th^ day. The actual events of negative RT-PCR were more in the AYUSH 64 group. The difference in two groups seems to be clinically significant though statistically it is not significant [p=0.28]. **(Table 2)**

**Table 2:** shows the RT-PCR result of 5^th^ day.

| **On 5<sup>th</sup> day** | **Positive (n %)** | **Negative (n %)** |
| --- | --- | --- |
| **Group 1 (AYUSH 64)** (n = 30) | 9(30) | 21(70) |
| **Group 2 (Control)** (n = 30) | 14(46.6) | 16(53.3) |
| $p=0.28$, fisher's exact test, two tailed. | | |

There was no statistically significant difference between two groups for fever and respiratory symptoms and lab parameters shows both groups were equivalent with each other. No serious adverse events reported from any group during the assessment period. **(Table 3, Table 4)**

## Discussion

This study was designed with the aim of the assessing the efficacy and safety of the Ayush- 64 in mild cases of COVID-19 when it was given in addition to the standard treatment. It was found that conversion of RT-PCR from positive to negative was not statistically different between AYUSH 64 and control group, though absolute event of conversion was more in AYUSH 64. No difference was observed for clinical and laboratory parameters between Ayush – 64 and control group. This no significant difference between both the groups may be because of less sample size or due to chance. A similar study using Ayush – 64 in addition with the standard treatment is now published as preprint and it shows significant beneficial effect of this intervention for COVID -19 when it was given with standard treatment in comparison to the placebo group. [22] In this study, two primary endpoints i.e mean duration of time to first day of clinical recovery was significantly less in intervention group as compared to the control group and proportion of patients with full clinical recovery was more in the intervention group as compared to the control. Quality of life score was also better in the Ayush – 64 group. There was no significant different in laboratory parameters. [22] This study has large sample size in comparison to our study and has better endpoints as compared to our study which relied heavily on RT-PCR. We have chosen the minimal sample size considering it as a new drug being explored in the disease first time and there was a plan to extend the study further if safety is established in the small sample size but it could not be done due to unavailability of the patients. In our study the primary endpoint was the conversion of RT-PCR from positive to negative. On this important criterion, Ayush 64 proved to be quite efficacious. Keeping this as an endpoint has its own pros and cons. As we wanted to avoid any subjectivity and bias due to open nature of the trial, we chosen RT-PCR conversion as the primary endpoint, but RT-PCR test results may be false positive as well as false negative and this may affect the overall analysis. [23] In our study there was no serious adverse event was reported while in the study by Chopra et al three serious adverse events were reported but these were from the control group and not from Ayush – 64 group. It shows that the drug has adequate safety in humans with the current dosing schedule. [22] Our study too reiterates the safety of Ayush 64.

COVID -19 is a disease associated with high morbidity and there is need of exploration of effective interventions to prevent and treat this disease on urgent basis which can be attempted if potential therapeutic interventions from modern medicines or complementary and alternative system are explored on fast-track basis. Our study was an attempt in this direction. Looking at the positive results in the study already published as preprint and encouraging even though statistically non-significant results in our study, the efficacy of Ayush -64 in the COVID-19 needs to be confirmed again in a large clinical trial in double blind manner with adequate sample size.

## Data Availability

Data is available with Principal Investigator - Dr. Pankaj Bhardwaj

## Acknowledgments

District Health Services of State of Rajasthan, Chief Medical Health Officer (CMHO) and his team, District administration are to be acknowledged in this present study.

